# Is metacarpophalangeal joint restriction an indicator of blood pressure?

**DOI:** 10.1101/2023.12.20.23300128

**Authors:** Rohan Shah, Sanat Phatak, Sarita Jadhav, Aboli Bhalerao, Shreya Mehra, Smita Dhadge, Jennifer Ingram, Chittaranjan Yajnik

## Abstract

Arterial wall stiffness is implicated in the etiopathogenesis of hypertension, reflecting medial stiffness and atherosclerotic plaques. In individuals with diabetes, musculoskeletal phenotypes like limited joint mobility (LJM) in the hands suggests soft tissue fibrosis and could mirror internal organ fibrosis including arterial stiffness.

In Quantifying hand stiffness, mean extension at the metacarpophalangeal (MCP) joint emerges as a promising metric. In a type 1 diabetes cohort, this correlated with structural fibrotic changes on MRI. The study introduces a clinically scalable methodology for measuring joint stiffness, with implications for community screening programs for hypertension.

In an ongoing study with 1885 participants, MCP extension showed a significant association with SBP (p <0.001). Linear regression, adjusted for age, gender, and diabetes status, reveals a negative correlation between average MCP extension and SBP. These pilot findings support earlier associations with an easier methodology. Future work involves establishing cutoffs in mean MCP extension for optimized screening algorithms, considering potential confounders like physical activity.

## Manuscript

Arterial wall stiffness plays a role in the etiopathogenesis of hypertension and owes to medial stiffness and atherosclerotic plaques.^1^ Tissue stiffness is believed to be a constitutional trait that could be phenotypically observable in other tissues outside the cardiovascular system. Joint stiffness and skin extensibility are potential non-invasive markers of tissue stiffness. In a healthy population of 96 school children, joint stiffness and skin extensibility were associated with both systolic and diastolic blood pressure (SBP and DBP).^2^ Joint stiffness was measured using individual Z scores of goniometry readings in five joints (shoulder, elbow, wrist, hip and ankle). DBP was reduced by 4.5 mmHg per standard deviation of joint mobility.

Joint mobility restriction occurs in the hands in both type 1 and type 2 diabetes, owing to inflammatory-fibrotic processes in various soft tissue structures on the palmar surface. The process may involve the palmar skin leading to flexion contractures (limited joint mobility, LJM); palmar fascia (Duypuytren’s disease, DD) and/ or flexor tendon sheaths (flexor tenosynovitis, FT/ trigger finger). We postulated that these musculoskeletal phenotypes may reflect internal organ fibrosis and stiffness-including arterial stiffness.^3^ In a Japanese cohort with type 2 diabetes, those with LJM (72 of 342 patients, diagnosed using a prayer sign or table test) had a higher carotid intima-media thickness (CIMT, 1.45 vs 1.14 mm) as well as a plaque score. LJM correlated with plaque score on multiple linear regression after adjusting for risk factors including age, duration of diabetes and smoking.^4^

The utility of using LJM as a clinical biomarker is hampered by a binary definition (present or absent) using a prayer sign, which could be subjective. In an attempt to bring quantitative rigour to hand mobility restriction, we saw that mean extension at the metacarpophalangeal (MCP) is a promising metric of palmar soft tissue stiffness and is affected not only by LJM but DD and FT as well. In our type 1 diabetes cohort, the MCP extension angle had structural fibrotic correlates on magnetic resonance imaging (MRI).^5^ The measurement can be performed using a protractor in the clinic. This contrasts with the methodology used by Uiterwaal et al which, while thorough, is time-consuming in practice, requires training, specialised equipment, and may be uncomfortable for large joint examinations.

In an ongoing study looking at the prevalence and severity of hand stiffness, we studied consecutive patients in the clinic with both type 1 and type 2 diabetes, and healthy subjects from a rural longitudinal cohort in Pune, India. We examined the hands for all these manifestations (LJM, DD, FT), hand grip strength and calculated mean MCP extension. Blood pressure was measured using a digital sphygmomanometer and an average of two readings was taken. Among 1885 study participants, 443 were on treatment for hypertension. Among 1442 individuals not on anti-hypertensive medication (770 [53.39%] male), 591 had diabetes (227 type 1, 364 type 2). The majority (n=800, 55.4%) had mean MCP angle extension between 30 and 60 degrees, while 123 (8.5%) had a range restricted to less than 30 degrees. This group had an average SBP that was 4.2 mm higher than in the 30-60 degrees extension group. (Table 1) They were also the oldest and had the highest prevalence of diabetes. An additional one-fifth of the group fulfilled Joint National Committee (JNC) 8 criteria for hypertension. Linear regression adjusted for age, gender and diabetes status showed a significant association between average MCP extension and SBP (Standardised β= -0.08; adjusted R^2^17.1%, p <0.001) but not DBP (Standardised β=-0.04, adjusted R^2^ 6.5%, p =0.10). These associations were held within the diabetic population when adjusted for gender, age and duration of diabetes. (Standardised β= -0.120, Adjusted R^2^ 19.2%, p=0.002 for SBP; Standardised β = -0.084, adjusted R^2^ 1.4% p =0.05, for DBP).

**Table 1.** Grouping by maximum passive metacarpophalyngeal joint extension and associated risk factors.

|  | MCP EXTENSION GROUP |  |  | p- value<br>(ANOVA) |
| --- | --- | --- | --- | --- |
|  | ≤ 30° (n=123) | 30-60° (n=800) | 60 - 90° (n=519) |  |
| Male n, % | 85.0 (69.1%) | 411.0 (51.4%) | 274.0 (52.8%) | - |
| Total Diabetes | 68.0 (55.3%) | 326.0 (40.8%) | 197.0 (38.1%) | - |
| Type 1 Diabetes | 20.0 (16.3%) | 113.0 (14.1%) | 94.0 (18.1%) | - |
| Age, years | 51.5 (37.5, 59.2) | 45.3 (27.8, 53.9) | 32.2 (26.6, 47.8) | .002 |
| Duration of Diabetes, years | 15.7 (6.9, 23.3) | 10.4 (4.1, 16.9) | 9.4 (4.6, 15.8) | .301 |
| Weight, kg | 64.6 (55, 73) | 61.8 (53.8, 71.7) | 63.1 (54.1, 71.9) | .628 |
| Height, cm | 162.9 (154.8, 170.3) | 160.4 (153.5, 168.5) | 163 (154, 170.5) | .055 |
| BMI | 24.4 (21.3, 27.3) | 24.1 (21.3, 27.1) | 24 (21.1, 26.8) | .817 |
| Systolic blood pressure, mm Hg | 120 (110, 136) | 115.8 (106, 128) | 113 (104, 122.5) | .020 |
| Diastolic blood pressure, mm Hg | 74.5 (66, 82) | 73 (65.5, 80) | 71.5 (65, 78) | .128 |
| Right-handgrip | 45.0 (31.7, 60) | 43.3 (35, 60) | 46.7 (36.7, 65.0) | .854 |
| Left-Handgrip | 45.0 (33.3, 60) | 43.3 (35, 60) | 46.7 (35.0, 65.0) | .807 |
| Newly Diagnosed Hypertension | 25.0 (20.3%) | 111.0 (13.9%) | 45.0 (8.7%) | <0.001 |

*Values are in n (%) or median (25<sup>th</sup> – 75<sup>th</sup> centile), p-value by ANOVA adjusted for age, gender, Duration of Diabetes, Participant type*

These pilot data from a large group of subjects support earlier reports of associations between blood pressure and joint stiffness.^2^ A simple, clinically scalable methodology for measuring joint stiffness promises translation into an additional indicator in community screening programs for hypertension, pending further validation. Our weaknesses include a cross-sectional analysis that precludes causal inference and a non-uniform distribution in age and diabetes. We acknowledge many unmeasured confounders such as physical work and exercise, that may add to joint stiffness. Future work would include establishing cutoffs in mean MCP extension as a step to optimise screening algorithms, leveraging a putative commonality between soft tissue and arterial stiffness.

## Data Availability

All data produced in the present study are available upon reasonable request to the authors

## References

1. Safar ME, Asmar R, Benetos A, Blacher J, Boutouyrie P, Lacolley P, Laurent S, London G, Pannier B, Protogerou A, et al. Interaction Between Hypertension and Arterial Stiffness. Hypertension. 2018;72(4):796–805. doi:10.1161/HYPERTENSIONAHA.118.11212

2. Uiterwaal CS, Grobbee DE, Sakkers RJ, Helders PJ, Bank RA, Engelbert RH. A relation between blood pressure and stiffness of joints and skin. Epidemiology.2003;14(2):223–227. doi:10.1097/01.EDE.0000040327.31385.9B

3. Phatak, S., Mahadevkar, P., Chaudhari, K. S., Chakladar, S., Jain, S., Dhadge, S., Jadhav, S., Shah, R., Bhalerao, A., Patil, A., Ingram, J. L., Goel, P., & Yajnik, C. S. (2023). Quantification of joint mobility limitation in adult type 1 diabetes. Frontiers in Endocrinology, 14. 10.3389/fendo.2023.1238825

4. Mineoka Y, Ishii M, Hashimoto Y, Tanaka M, Nakamura N, Katsumi Y, Isono M, Fukui M. Relationship between limited joint mobility of hand and carotid atherosclerosis in patients with type 2 diabetes. Diabetes Res Clin Pract. 2017;132:79–84. doi:10.1016/j.diabres.2017.07.002

5. Phatak S, Ingram JL, Goel P, Rath S, Yajnik C. Does hand stiffness reflect internal organ fibrosis in diabetes mellitus?. Front Clin Diabetes Healthc. 2023;4:1198782. doi:10.3389/fcdhc.2023.1198782

